# A survey and antibody test following the surge of SARS-CoV-2 Omicron infection in China

**DOI:** 10.1101/2023.02.28.23286535

**Authors:** Yichuan Yao, Yunru Yang, Qiqin Wu, Mengyao Liu, Wei Bao, Qiutong Wang, Meijun Cheng, Yunuo Chen, Yuan Cai, Mei Zhang, Jingxue Yao, Hongliang He, Changjiang Jin, Tian Xue, Changcheng Zheng, Tengchuan Jin, Dali Tong

## Abstract

The surge of SARS-CoV-2 Omicron infection in most Chinese residents at the end of 2022 provided a unique opportunity to understand how the immune system responds to the Omicron infection in a population with limited contact to prior SARS-CoV-2 variants. Moreover, whether the prototype SARS-CoV-2 booster vaccination could help induce the antibody against Omicron variants? Here, we tested the level of IgG, IgA, and IgM specific to the prototype SARS-CoV-2 spike RBD (Receptor Binding Domain) from the collected blood samples from 636 individuals. Sequential inoculation of different vaccines showed higher IgG levels after infection. As the antibody level against Omicron BA.5, BF.7, and XBB 1.5 of the individuals has highly positive correlation with the antibody level against prototype SARS-CoV2, the IgG level specific to the prototype SARS-CoV-2 spike RBD could also represent the IgG level against Omicron variants. Furthermore, the 4^th^ booster vaccination could induce a comparable antibody level against prototype, Omicron BA.5, BF.7, and XBB 1.5 variants in the patients with 2 or 3-dose vaccination and protect people from being infected. In conclusion, these data suggest that the prototype SARS-CoV-2 booster vaccination helps induce a high level of antibody against prototype, BA.5, BF.7, and XBB 1.5 variants after Omicron infection.

---

Coronavirus disease 2019 (COVID-19), caused by a severe acute respiratory syndrome coronavirus 2 (SARS-CoV-2) infection, continues to spread rapidly across the globe and threaten global public health. Since the spread of SARS-CoV-2, it has developed into a lot of variants, such as Alpha, Beta, Delta, Omicron, etc. Currently, the Omicron variant has become the major circulating virus strain, including the Omicron offshoot BA.5, BF.7, XBB and so on. More than 30 mutations in the spike (S) protein and 15 mutations in the RBD of the S protein were found in the Omicron variant, which induced the escape neutralization activity of the Omicron variant by most of the identified anti-SARS-CoV-2 neutralization antibodies^1,2^.

Many types of vaccines have been developed to control the infection and spread of SARS-CoV-2, including vaccines based on messenger RNA (mRNA)^3^, viral vectors^4,5^, recombinant proteins^6^, and inactivated SARS-CoV-2^7^. Immunization with these vaccines showed a reduction in infection rates and post-infection mortality. A booster dose of inactivated SARS-CoV-2 vaccine led to a significant increase in a neutralizing immune response against prototype SARS-CoV-2 and the Omicron variant with incomplete escape^8^. On the other hand, repeated vaccination with inactivated virus vaccine may recall a strong immune response to target the prototype strain because of the immune imprint, while it also inhibits immune responses to new Omicron variants even with the Omicron BA.5 vaccine boost^1,9^. These reports suggest that current herd immunity may not efficiently prevent the infection of the highly mutagenic Omicron variants.

From mid-December of 2022 to early January of 2023, the vast majority of Chinese residents have experienced a surge of SARS-CoV-2 Omicron infection. According to data released by the Chinese Center for Disease Control and Prevention, the SARS-CoV-2 that caused the spread of this current epidemic is mainly Omicron BA.5 and BF.7 variant. Therefore, we conducted a survey 750 people for SARS-CoV-2 infection and collected blood samples from 636 people in Hefei, Anhui Province, China. The demographic and epidemiological characteristics of the 636 people were summarized in Table 1. With regard to the people surveyed, the median age was 32 years (interquartile range: 27 and 49 years; range:2 to 69 years). 22 people were younger than 15 years and 57 people were older than 60 years. 308 of them are women. Among the 636 people, 441 people were infected as confirmed by antigen or nucleic acid testing, 142 people were negative by both testings, with the other 53 people have not been tested by the time of blood collection. The ratio of being tested positive was similar between men and women. The percentage of infected patients increased with age (Spearman Correlation Analysis, r = 1, P = 0.0167). 611 people (96.07%) are fully vaccinated: 2-dose inactivated SARS-CoV2 vaccine (10.53%); 3-dose recombinant protein vaccines (4.09%); 2-dose inactivated SARS-CoV2 vaccine with 1-dose inactivated SARS-CoV2 vaccine booster (57.55%); 2-dose inactivated SARS-CoV2 vaccine with 1-dose recombinant protein vaccine booster (12.42%); 2-dose inactivated SARS-CoV2 vaccine with 1-dose AdVs vaccine booster (1.26%); 2-dose mRNA vaccines (0.63%); 3-dose mRNA vaccines (0.16%); 4-dose vaccines (8.49%). 19 people (2.99%) were unvaccinated; 4 people (0.63%) were vaccinated with 1-dose inactivated SARS-CoV2 vaccine and 2 (0.32%) were vaccinated with 1-dose recombinant protein SARS-CoV2 vaccine. Of the 611 fully vaccinated people, 5 people (0.86%) received the latest vaccine less than 7 days; 11 people (1.80%) received the latest vaccine between 7 to 13 days; 49 people (8.02%) received the latest vaccine between 14 to 30 days; 6 people (0.98%) received the latest vaccine between 1 to 6 months; and the others (540, 88.38%) received the latest vaccine more than 6 months. The infected percentage was 39.13% in the group that received the last dose vaccine less than 30 days, which is lower than the infected percentage (75.82%) of the group that received the last dose vaccine more than 1 month ago. The result suggested that a boost dose vaccine received in might protect the people from SARS-CoV-2 infection.

**Table 1.** Demographics and baseline characteristics of the uninfected people and patients infected with SARS-CoV2 Omicron variants.

| Characteristics | No. (%) of people <sup>a</sup> |
| --- | --- |
| **Age groups, years** |  |
| 0-15 | 22 (3.46%) |
| 16-30 | 206 (32.39%) |
| 31-45 | 203 (31.92%) |
| 46-60 | 148 (23.27%) |
| >60 | 57 (8.96%) |
| **Sex** |  |
| Male | 328 (51.57%) |
| Female | 308 (48.43%) |
| **Confirmed with SARS-CoV2 infection <sup>b</sup>** |  |
| Uninfected | 142 (22.33%) |
| Infected | 441 (69.34%) |
| Not sure | 53 (8.33%) |
| **Infection of male** |  |
| Uninfected | 74 (22.56%) |
| Infected | 224 (68.29%) |
| Not sure | 30 (9.15%) |
| **Infection of female** |  |
| Uninfected | 68 (22.08%) |
| Infected | 217 (70.45%) |
| Not sure | 23 (7.47%) |
| **Age** |  |
| 0-15 | 8 (36.36%) |
| 16-30 | 137 (66.50%) |
| 31-45 | 139 (68.47%) |
| 46-60 | 112 (75.68%) |
| >60 | 45 (78.95%) |
| **SARS-CoV-2 vaccination strategy <sup>c</sup>** |  |
| Unvaccinated | 19 (2.99%) |
| 1-dose IV | 6 (0.94%) |
| 2-dose IV | 67 (10.53%) |
| 3-dose IV | 366 (57.55%) |
| 3-dose RP | 26 (4.09%) |
| 2-dose IV & 1-dose RP | 79 (12.42%) |
| 2-dose mRNA | 4 (0.63%) |
| 3-dose mRNA | 1 (0.16%) |
| 2-dose IV & 1-dose Adv | 8 (1.26%) |
| 4-dose vaccine | 54 (8.49%) |

| Interval days of latest SARS-CoV-2 vaccination and diagnosis day |  |
| --- | --- |
| 0-6 Days | 0/5 (0%) |
| 7-13 Days | 0/11 (0%) |
| 14-30 Days | 27 (55%) |
| 1-6 Month | 6 (100%) |
| >6 Month | 408 (76%) |

- 1 a. Data are shown as number (%) or number/total number (%) in the 0-6 days and 7- - 2 13 days of the interval days of latest SARS-CoV-2 vaccination and diagnosis day. - 3 b. Infected: antigen or nucleic acid testing positive; uninfected: antigen or nucleic acid - 4 testing negative; Not sure: no antigen or nucleic acid test. - 5 c. IV: Inactivated SARS-CoV-2 Virus vaccine; RP: Recombinant Protein vaccine; - 6 mRNA: mRNA vaccine; Adv: Adv-based vaccine.

The clinical characteristics of the 441 individuals who tested positive for antigen or nucleic acid are summarized in Table 2. Only 4 patients (0.91%) reported no special symptom, and the most common symptoms were fever (86.62%), cough (84.6%), weakness (67.35%), sputum production (65.99%), headache (50.79%), myalgia (50.79%), sore throat (49.43%), runny nose (35.37%), loss of taste and smell (30.39%) and conjunctivitis (3.17%). The mean duration of symptoms is 5 days (range, 1 to 20 days). 24.26% of patients felt mild symptoms while 12.70% experienced severe symptoms. 355 patients (80.50%) needed medication to relieve symptoms. Drugs included Ibuprofen (93.80%), Paracetamol (44.79%), Chinese medicine (44.79%), Antibiotic (2.82%) and Paxlovid (0.56%).

**Table 2.** Clinical features of the 441 patients infected with SARS-CoV2 Omicron variants.

| Characteristics | No. (%) of patients <sup>a</sup> |
| --- | --- |
| **Initial presenting symptoms** |  |
| Fever | 382 (86.62%) |
| Cough | 373 (84.6%) |
| Weakness | 297 (67.35%) |
| Sputum production | 291 (65.99%) |
| Headache | 224 (50.79%) |
| Myalgia | 224 (50.79%) |
| Sore throat | 218 (49.43%) |
| Runny nose | 156 (35.37%) |
| Loss of taste and smell | 134 (30.39%) |
| Conjunctivitis | 14 (3.17%) |
| **Symptom Score <sup>b</sup>** |  |
| 0-3 | 107 (24.26%) |
| 4-7 | 278 (63.04%) |
| 8-10 | 56 (12.7%) |
| **Medicine-use** |  |
| Medicine | 355 (80.50%) |
| Non-medicine | 86 (19.50%) |
| **Medicine type <sup>c</sup>** |  |
| Ibuprofen | 333 (93.80%) |
| Paracetamol | 159 (44.79%) |
| Chinese medicine | 159 (44.79%) |
| Antibiotic | 10 (2.82%) |
| Paxlovid | 2 (0.56%) |

- 3 a. Data are shown as number (%) - 4 b. Symptom Score was provided by patients, 0: feel no special symptom. 10: feel 5 severest symptom. No patient with severe or critically ill COVID-19 diagnosed by 6 the hospital was involved in the survey. - 7 c. The patients who took this kind of medicine were summarized in the table below, 8 and they may take other kinds of medicine at the same time.

Then, we tested the antibody level, including IgG, IgA, and IgM, of prototype SARS-CoV-2 by a set of chemical luminescence kits that can quantitatively and sensitively measure the levels of IgG, IgA, and IgM specific to the prototype SARS-CoV-2 spike RBD^10,11^. We found that the IgG of infected patients was higher than that of uninfected people (Figure 1 A). Besides, IgA and IgM levels were low in both the infected patients and the uninfected people, which was lower than the cut-off of the positive control (Figure S1 A and B)^10^. As a result, we focus on the analysis of IgG levels. There was no significant change in the IgG level of the patients with different ages, sex, or BMI (Body Mass Index, Figure S1 C-E). The IgG level in patients with severe symptoms is higher than the patients with mild symptoms (Figure 1 B). A similar situation is that the IgG level of patients taking medicines is higher than that of patients with non-drug treatment (Figure 1 C), which may be due to the worse symptoms of the patients who taking medicine (Figure S1 F). No significant changes of IgG were found in the groups with different types of medicines (Figure S2 G).

**Figure 1.**
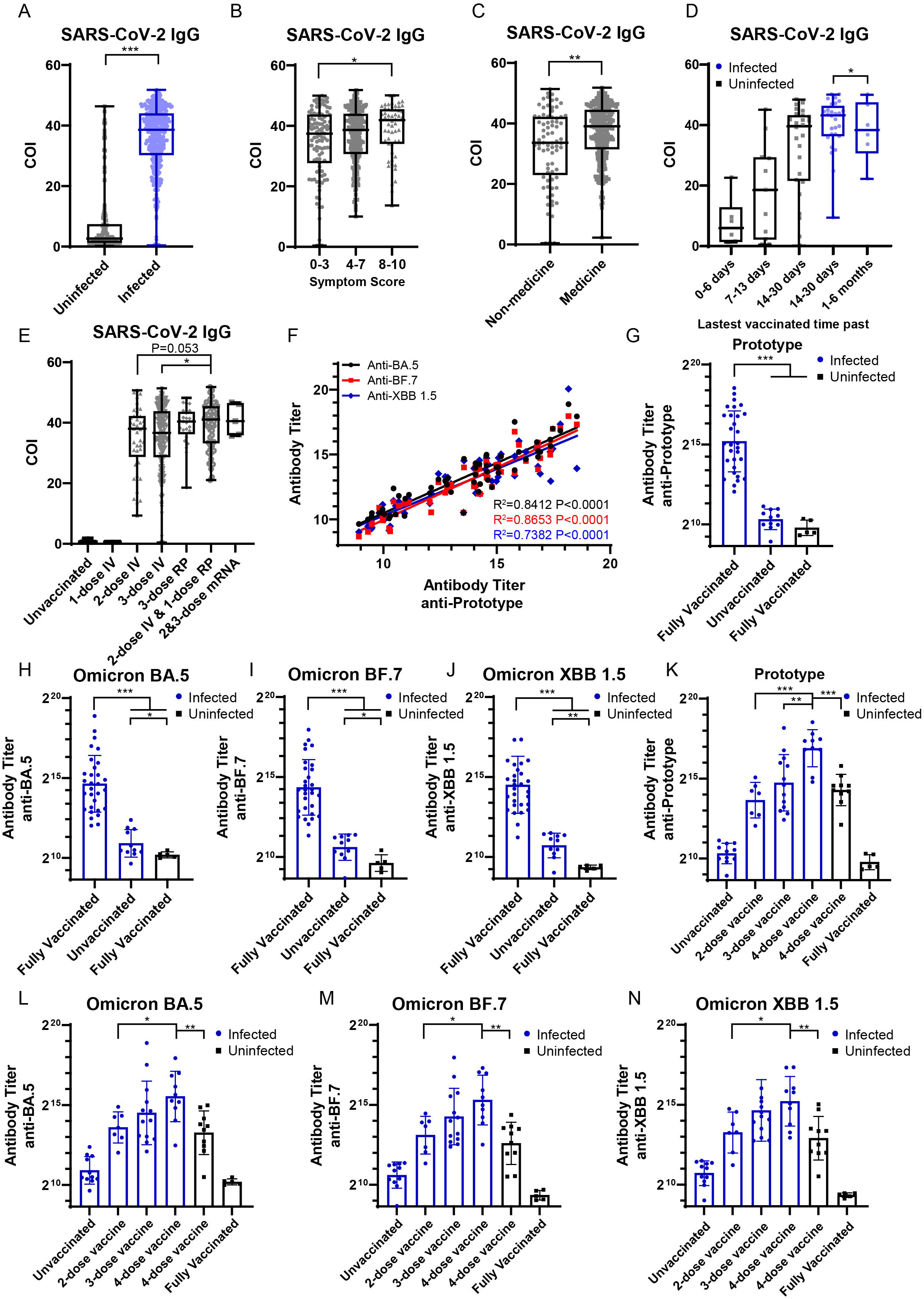
The booster vaccination of prototype vaccines could help patients produce higher IgG antibody against the Omicron variants. A. The IgG level against prototype SARS-CoV2 of the infected and uninfected people tested by chemical luminescence kits. (n = 108 in the uninfected group; n = 441 in the infected group) B. The IgG level of infected patients with different symptom score. (n = 107 in the 0-3 group; n = 278 in the 4-7 group; n = 56 in the 8-10 group) C. The IgG level of infected patients who take medicine or not. (n = 355 in the medicine group; n = 86 in the non-medicine group) D. The IgG level of infected and uninfected people who received the lasted vaccine in 6 days, 7-13 days, 13-30 days and 1-6 months. (n = 5 in the 0-6 uninfected days; n = 11 in the 7-13 days uninfected group; n = 22 in the 14-30 days uninfected group; n =27 in the 14-30 days infected group; n = 6 in the 1-6 months infected group) E. The IgG level of infected patients with different vaccination strategy. (IV: Inactivated SARS-CoV-2 Virus vaccine; RP: Recombinant Protein vaccine; n = 19 in the unvaccinated group; n = 6 in the 1-dose IV group; n = 67 in the 2-dose IV group; n = 366 in the 3-dose IV group; n = 26 in the 3-dose RP group; n = 79 in the 2-dose IV & 1-dose RP group; n = 5 in the mRNA group. F. The Pearson correlation coefficient analysis of antibody titer against prototype SARS-CoV2 and Omicron variants. (n = 56) G. Quantitative analysis of RBD antibody titer against prototype SARS-CoV2 calculated by ELISA. (n = 30 in the fully vaccinated infected group; n = 11 in the unvaccinated infected group; n = 5 in the fully vaccinated uninfected group) H. Quantitative analysis of RBD antibody titer against Omicron BA.5 variant calculated by ELISA. (n = 30 in the fully vaccinated infected group; n = 11 in the unvaccinated infected group; n = 5 in the fully vaccinated uninfected group) I. Quantitative analysis of RBD antibody titer against Omicron BF.7 variant calculated by ELISA. (n = 30 in the fully vaccinated infected group; n = 11 in the unvaccinated infected group; n = 5 in the fully vaccinated uninfected group) J. Quantitative analysis of RBD antibody titer against Omicron XBB 1.5 variant calculated by ELISA. (n = 30 in the fully vaccinated infected group; n = 11 in the unvaccinated infected group; n = 5 in the fully vaccinated uninfected group) K. Quantitative analysis of RBD antibody titer against prototype SARS-CoV2 calculated by ELISA. (n = 11 in the unvaccinated infected group; n = 7 in the 2-dose infected group; n = 13 in the 3-dose infected group; n = 10 in the 4-dose infected group; n = 10 in the 4-dose uninfected group; n = 5 in the fully vaccinated uninfected group) L. Quantitative analysis of RBD antibody titer against Omicron BA.5 variant calculated by ELISA. (n = 11 in the unvaccinated infected group; n = 7 in the 2-dose infected group; n = 13 in the 3-dose infected group; n = 10 in the 4-dose infected group; n = 10 in the 4-dose uninfected group; n = 5 in the fully vaccinated uninfected group) M. Quantitative analysis of RBD antibody titer against Omicron BF.7 variant calculated by ELISA. (n = 11 in the unvaccinated infected group; n = 7 in the 2-dose infected group; n = 13 in the 3-dose infected group; n = 10 in the 4-dose infected group; n = 10 in the 4-dose uninfected group; n = 5 in the fully vaccinated uninfected group) N. Quantitative analysis of RBD antibody titer against Omicron XBB 1.5 variant calculated by ELISA. (n = 11 in the unvaccinated infected group; n = 7 in the 2-dose infected group; n = 13 in the 3-dose infected group; n = 10 in the 4-dose infected group; n = 10 in the 4-dose uninfected group; n = 5 in the fully vaccinated uninfected group) Values are Median (Min to Max) or geometric mean ± geometric standard deviation for antibody titer. Mann-Whitney test, *: P < 0.05; **: P < 0.01; ***: P < 0.001.

Vaccination is one of the most effective ways to control the spread of SARS-CoV-2. Therefore, we analyzed the IgG levels of people who received the last dose vaccine in different days. The IgG level increased with the time passing by in 30 days (One-way ANOVA of uninfected group, IgG: P = 0.0005) and the infection would induce higher IgG level (Figure 1 D). In addition, the vaccination strategy caused different IgG after the Omicron infection. Omicron infection failed to induce measurable IgG levels among those unvaccinated and 1-dose inactivated SARS-CoV-2 vaccinated individuals. Among the fully vaccinated groups, the 2-dose inactivated SARS-CoV2 vaccine with dose recombinant protein vaccine booster group showed a higher IgG level than the dose inactivated SARS-CoV2 vaccine group (P = 0.053) and the 3-dose inactivated SARS-CoV2 vaccine group (P = 0.026, Figure 1 E).

The prototype SARS-CoV-2 booster vaccination shows a high ability to induce the IgG antibody against prototype SARS-CoV2, while how the prototype SARS-CoV-2 booster vaccination help protect the Omicron infection remains unknown. To test the antibody level against different variants of SARS-CoV2, we collected the plasma of 41 infected patients (11 unvaccinated patients, 7 with 2-dose inactivated SARS-CoV2 vaccine, 13 with 3-dose inactivated SARS-CoV2 or recombinant protein vaccine and 10 with 4-dose inactivated SARS-CoV2 or recombinant protein vaccine) and 15 uninfected people, including 10 people received the latest boost (4^th^ dose) less than 3 months and 5 people received the latest boost more than 6 months. We then tested the antibody titer (IgG) in the plasma against the prototype, Omicron BA.5, BF.7, and XBB 1.5 variants using ELISA test^4^. Surprisingly, we found that the antibody level against Omicron BA.5, BF.7, and XBB 1.5 of the individuals has a high positive correlation with the antibody level against prototype SARS-CoV2 (Figure 1 F). These data suggests that the antibody level against prototype SARS-CoV2 could represent the antibody level against the Omicron BA.5, BF.7, and XBB 1.5 of both infected and uninfected people. The results also showed that the infection of Omicron induced a high level of antibody against prototype, Omicron BA.5, BF.7, and XBB 1.5 variants in fully vaccinated patients, which was consistent to the result of chemical luminescence kits. On the other hand, the antibody against Omicron BA.5, BF.7, and XBB 1.5 variants induced by Omicron infection of unvaccinated group was higher than the uninfected fully vaccinated group but not the antibody against prototype SARS-CoV2 (Figure 1 G-J). We also found that the 4^th^ booster vaccination could induce comparable antibody level against prototype, Omicron BA.5, BF.7, and XBB 1.5 variants with the 2 or 3-dose infected patients and induced higher antibody level against prototype, Omicron BA.5, BF.7, and XBB 1.5 variants after infection (Figure 1 K-N).

In conclusion, in this study with blood samples collected from 636 people in Hefei, Anhui Province, China in middle of January 2023, around 3 weeks after the quick pandemic infection, we found the last booster vaccine received could prevent the people from being infected with SARS-CoV-2. The IgG level of patients with severe symptoms is higher than the patients with mild symptoms. Moreover, the vaccination strategy resulted in different IgG level after the Omicron infection. The Omicron infection could not induce the IgG level in the unvaccinated and 1-dose inactivated SARS-CoV-2 patients. The antibody level against Omicron BA.5, BF.7, and XBB 1.5 of the individuals has a high positive correlation with the antibody level against prototype SARS-CoV2. In addition, the 4^th^ booster vaccination could induce the comparable antibody level against prototype, Omicron BA.5, BF.7, and XBB 1.5 variants with 2 or 3-dose infected patients and induce higher antibody level after infection. These data suggest that booster vaccination of prototype vaccines could help patients produce higher IgG antibody against the Omicron variants even though it recalls a strong immune response to target the prototype strain because of the immune imprint.

## Data Availability

All data produced in the present study are available upon reasonable request to the authors.
All data produced in the present work are contained in the manuscript.

## Acknowledgments

We thank all colleagues from the First Affiliated Hospital of USTC and the Hospital of USTC, for their support during the study. We thank all of the people who take part in the project for the survey and the blood collection.

## Ethics approval

All procedures were conducted in accordance with the Principles for the Medical Ethical Committee of the First Affiliated Hospital of USTC (approval number: 2023-ky-001).

## Authors’ contributions

D.T., T.J., C.Z., T.X., C.J., and H.H. conceived the project and designed the experiments. D.T., Y. Yao. and Y. Yang. designed and analyzed the questionnaire, tested antibody titer, and wrote the manuscript. Q.Wu. and M. L. did chemical luminescence kits, W. B. analyzed the questionnaire. Q. Wang Collected the blood from the people. M. C., Y. Chen., Y. Cai., M. Z., J. Y., H. H., and C. J. worked on data collection, analysis, and discussion. All authors edited and proofread the manuscript.

## Reference

1 Cao, Y. et al. Omicron escapes the majority of existing SARS-CoV-2 neutralizing antibodies. Nature 602, 657–663, doi:10.1038/s41586-021-04385-3 (2022).

2 Wang, Q. et al. Antibody evasion by SARS-CoV-2 Omicron subvariants BA.2.12.1, BA.4 and BA.5. Nature 608, 603–608, doi:10.1038/s41586-022-05053-w (2022).

3 Polack, F. P. et al. Safety and Efficacy of the BNT162b2 mRNA Covid-19 Vaccine. N Engl J Med 383, 2603–2615, doi:10.1056/NEJMoa2034577 (2020).

4 Tong, D. et al. Single-dose AAV-based vaccine induces a high level of neutralizing antibodies against SARS-CoV-2 in rhesus macaques. Protein & Cell 14, 69–73, doi:10.1093/procel/pwac020 (2022).

5 Zhu, F. C. et al. Immunogenicity and safety of a recombinant adenovirus type-5-vectored COVID-19 vaccine in healthy adults aged 18 years or older: a randomised, double-blind, placebo-controlled, phase 2 trial. Lancet 396, 479–488, doi:10.1016/S0140-6736(20)31605-6 (2020).

6 Yang, J. et al. A vaccine targeting the RBD of the S protein of SARS-CoV-2 induces protective immunity. Nature 586, 572–577, doi:10.1038/s41586-020-2599-8 (2020).

7 Zhang, Y. et al. Safety, tolerability, and immunogenicity of an inactivated SARS-CoV-2 vaccine in healthy adults aged 18-59 years: a randomised, double-blind, placebo-controlled, phase 1/2 clinical trial. Lancet Infect Dis 21, 181–192, doi:10.1016/S1473-3099(20)30843-4 (2021).

8 Yu, X. et al. Reduced sensitivity of SARS-CoV-2 Omicron variant to antibody neutralization elicited by booster vaccination. Cell Discov 8, 4, doi:10.1038/s41421-022-00375-5 (2022).

9 Gao, B. et al. Repeated vaccination of inactivated SARS-CoV-2 vaccine dampens neutralizing antibodies against Omicron variants in breakthrough infection. Cell Res, 1–4, doi:10.1038/s41422-023-00781-8 (2023).

10 Ma, H. et al. Serum IgA, IgM, and IgG responses in COVID-19. Cell Mol Immunol 17, 773–775, doi:10.1038/s41423-020-0474-z (2020).

11 Ma, H. et al. Decline of SARS-CoV-2-specific IgG, IgM and IgA in convalescent COVID-19 patients within 100 days after hospital discharge. Sci China Life Sci 64, 482–485, doi:10.1007/s11427-020-1805-0 (2021).

