## Supplemental File for "A survey and antibody test following the surge of SARS-CoV-2 Omicron infection in China"

### Materials and Methods

#### Survey and Human blood samples

This study was reviewed and approved by the Medical Ethical Committee of the First Affiliated Hospital of USTC (approval number: 2023-ky-001). The surveys were collected from 750 people during the Omicron Variants pandemic from December 2022 to January 2023 in Hefei, Anhui Province, China. Blood samples were from 636 people at the Hospital of USTC. No patients with severe or critically ill COVID-19 diagnosed by the hospital were involved in the survey. The history of vaccination was recorded and the patients were categorized as unvaccinated, 2-dose vaccine, 3-dose vaccine, and 4-dose vaccine.

#### Protein expression and purification

The methods for purifying the SARS-CoV-2 RBD [amino acid (AA) 321–591], SARS-CoV-2 RBD variants, and human ACE2 extracellular domain (AA 19 to 615) followed previous research. In brief, target genes were inserted into the pTT5 vector, which contains a IFNA1 signal peptide at the N-terminus and a TEV enzyme site connected to the human IgG1 Fc at the C-terminus. The expression vectors were then transfected transiently into HEK-293F cells using polyethyleneimine (Polyscience). After 3 days, the supernatant was collected by centrifugation at 5 000 ×*g* for 15 min at 4 °C. About ¼ volume of 1 × PBS was added to adjust the pH of the supernatant. The supernatant was then loaded onto the protein A column and the target protein was eluted with 0.1

M acetic acid on ÄKTA pure (GE Healthcare). The collected protein was added to 1 mM edetate disodium (EDTA), 5 mM dithiothreitol (DTT), and tobacco etch virus (TEV) protease to remove Fc on a shaker in a 4 °C freezer. After dialysis in 1 × PBS, tandem protein A and nickel columns were used for further purification. Both Fc and undigested protein were loaded onto the protein A column and TEV (6 × His tag) was loaded onto the nickel column. The target protein was collected during flow-through.

**ELISA**

Nunc MaxiSorp plates were coated with 3 μg/mL recombinant RBD, BA.5 RBD, BF.7 RBD or XBB 1.5 RBD at room temperature 2 h. After washing four times with PBS (3 min each time), the plates were blocked with 5% non-fat milk in PBS at room temperature for 2 h. Serially diluted serum (5% non-fat milk in PBST (PBS with 0.1% Tween-20) was added to the plates, which were then incubated at room temperature for 1 h. After washing three times with PBST (3 min each time), horseradish peroxidase (HRP)-conjugated goat anti-human IgG were added, followed by incubation at room temperature for 1 h. After washing three times with PBST (3 min each time), TMB substrate was added for 8 min, then stopped by 1 M $H_2SO_4$. Absorbance at 450 nm was measured with a microplate reader. Antibody titer was calculated as the dilution of the serum that induced an A450 value twice that of the A450 value of the negative control.

**Statistical analyses**

All data are presented as Median (Min to Max) or geometric mean ± geometric standard deviation for antibody titer. Mann-Whitney test, One-way analysis of variance

(ANOVA), the Pearson Correlation Analysis and Spearman Correlation Analysis were used in the test. Quantification graphs were analyzed using GraphPad Prism v8 (GraphPad Software). *: $P < 0.05$; **: $P < 0.01$; ***: $P < 0.001$.

Figure. S1

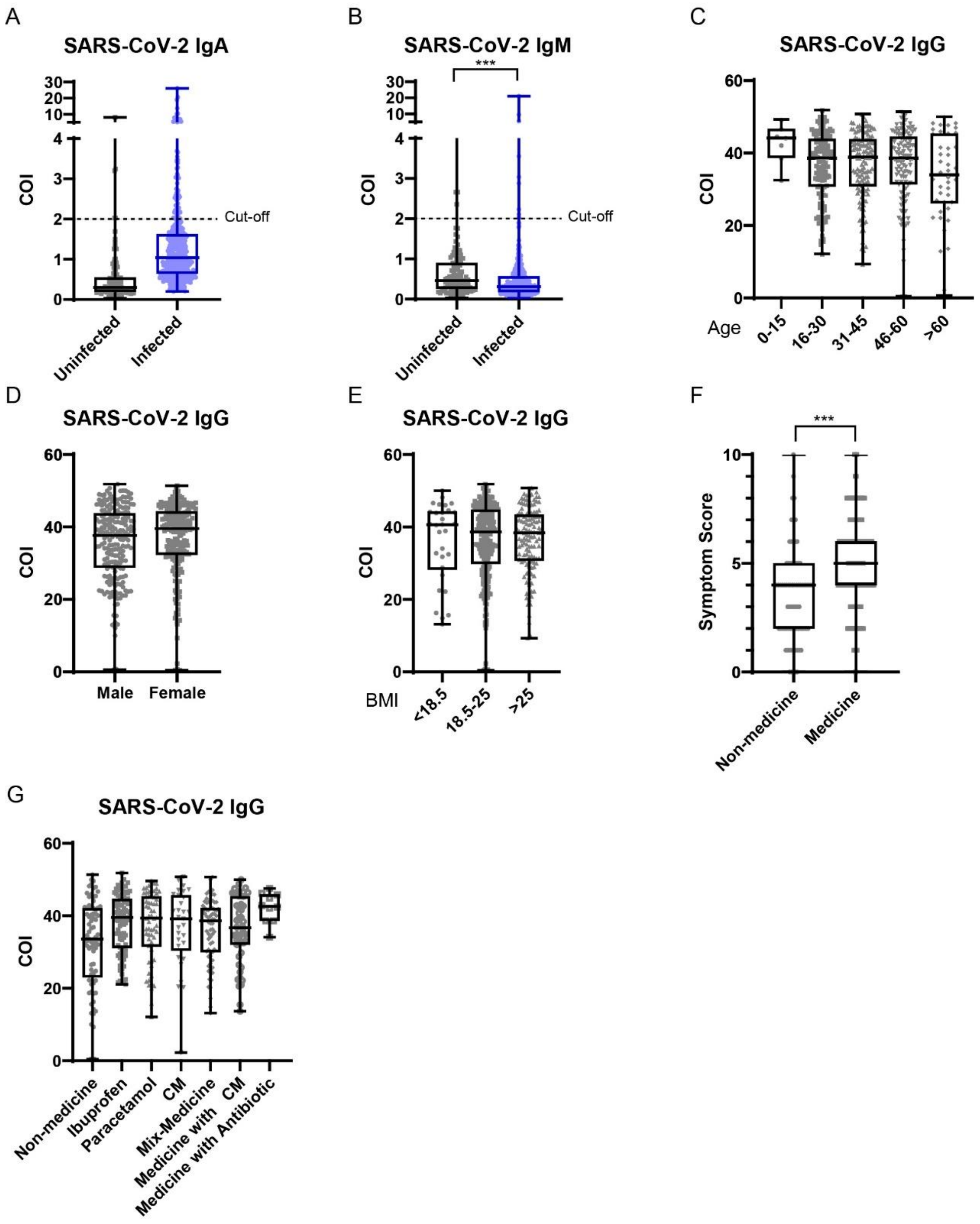


Figure. S1 No significant change in the IgG level of the patients with different ages, sex, or BMI.

A. The IgA level against prototype SARS-CoV2 of the infected and uninfected people tested by chemical luminescence kits. (n = 108 in the uninfected group; n = 441 in the infected group; Cut-off: the COI of IgA with positive control)

B. The IgM level against prototype SARS-CoV2 of the infected and uninfected people tested by chemical luminescence kits. (n = 108 in the uninfected group; n = 441 in the infected group; Cut-off: the COI of IgM with positive control)

C. The IgG level of infected patients with different age. (n = 8 in the 0-15 group; n = 137 in the 16-30 group; n = 139 in the 31-45 group; n = 112 in the 46-60 group; n = 45 in the >60 group)

D. The IgG level of infected patients with different sex. (Male: n = 224; Female: n = 217)

E. The IgG level of infected patients with different BMI. (BMI < 18.5: n = 30; BMI 18.5 - 25: n = 276; BMI > 25: n = 135)

F. The symptom score of infected patients who take medicine or not. (n = 355 in the medicine group; n = 86 in the non-medicine group)

G. The IgG level of infected patients who take different medicine. (CM: Chinese Medicine; n = 86 in the non-medicine group; n = 89 in the Ibuprofen only group; n = 62 in the Paracetamol only group; n = 32 in the Chinese Medicine only group; n = 68 in the Ibuprofen and Paracetamol group; n = 51 in the Chinese Medicine and Ibuprofen or Paracetamol group; n = 10 in the Medicine with Antibiotic and Ibuprofen or Paracetamol group; the other kind of medicine was not analyzed)
